# Outcome Orientation vs Problem Orientation: Preliminary Validation of a Novel Cognitive Assessment Tool and Its Relationship to Burnout in Advanced Practice Providers

**DOI:** 10.64898/2026.02.20.26346714

**Authors:** Ben Cartner, Stacy Schmauss, Matthew Bucala, Michael Y. Ghim, James Guerrini

**Affiliations:** Wake Forest School of Medicine, Wake Forest University, Winston-Salem, NC; Department of Transplant Surgery, University of Michigan, Ann Arbor, MI; Emergency Medicine and Urgent Care, Atrium Health Wake Forest Baptist, Winston-Salem, NC; Urgent Care, Atrium Health Wake Forest Baptist, Winston-Salem, NC

**Author notes:** Corresponding Author: Ben Cartner, MMS, MS, PA-C Wake Forest School of Medicine 475 Vine St, Winston-Salem, NC 27101.

**Keywords:** Burnout, Professional Fulfillment, Advanced Practice Providers, Physician Assistants, Nurse Practitioners, Emergency Medicine, Urgent Care, Cognitive Assessment

## Abstract

**Background:** Advanced Practice Providers (APPs) in emergency and urgent care settings experience high burnout rates, yet limited research examines cognitive factors influencing professional fulfillment. The Empowerment Dynamic framework suggests outcome-oriented thinking may protect against burnout compared to problem-oriented patterns.

**Objective:** To examine relationships between cognitive mindset orientation, professional fulfillment, and burnout among APPs while providing preliminary validation of a novel cognitive assessment instrument.

**Methods:** Cross-sectional survey of licensed APPs working in emergency departments and urgent care facilities across two health systems (July–October 2025). Professional fulfillment and burnout were measured using the Stanford Professional Fulfillment Index; cognitive orientation was assessed using a newly developed 22-item instrument.

**Results:** Among 98 respondents (19.5% response rate), mean professional fulfillment was 5.8 and mean burnout was 4.5; 40.8% met burnout criteria. Professional fulfillment and burnout were inversely correlated (r = −0.62; P < .001). Problem orientation correlated positively with burnout (r = 0.56) and negatively with fulfillment (r = −0.36), while outcome orientation showed opposite patterns (burnout: r = −0.57; fulfillment: r = 0.44). In multivariable models, outcome orientation remained independently associated with lower burnout (β = −1.51; P = .003) and higher fulfillment (β = 1.73; P = .002).

**Conclusions:** Cognitive mindset orientation is associated with burnout and professional fulfillment among APPs. The novel assessment instrument demonstrates acceptable psychometric properties. Future longitudinal studies are needed to establish causality and evaluate cognitive interventions for burnout prevention.

## INTRODUCTION

Healthcare worker burnout has reached epidemic proportions, with significant implications for provider well-being, patient safety, and healthcare sustainability.^1^ The International Classification of Diseases (ICD-11) defines burnout as a syndrome resulting from chronic workplace stress, characterized by exhaustion, cynicism, and reduced professional efficacy.^2^ However, the field of burnout research remains a shifting landscape, with findings largely dependent on how studies define and operationalize key terms.^3,4^ National surveys indicate approximately 50% of physicians experience burnout symptoms, though rates vary considerably based on measurement approaches and specialty.^5,6^

While extensive research has examined physician burnout, substantially less attention has been directed toward Advanced Practice Providers (APPs), including Physician Assistants (PAs) and Nurse Practitioners (NPs).^7–9^ This knowledge gap is further complicated by evolving national assessment strategies. Drawing on the Professional Fulfillment Index (PFI), 2018 AAPA data established baseline rates for emergency medicine PAs: 34.5% burnout, 50.8% work exhaustion, 21.5% interpersonal disengagement, and 72.3% professional fulfillment.^10^ In parallel, the NCCPA began tracking specialty-specific burnout and job satisfaction in 2020, with 30.6% of emergency medicine PAs (including urgent care) reporting at least one symptom of burnout; by 2024, emergency medicine PAs had the highest burnout rate (40.6%), and the 2023 survey identified urgent care as a distinct specialty with 39.4% burnout.^11,12,13^ More recent AAPA Morale and Burnout data indicate that the proportion of emergency medicine PAs meeting the PFI burnout threshold ranged from approximately 44% in 2020 to 58% in 2023 and 57% in 2024, consistently exceeding the overall PA average.^14^ In this context, it is noteworthy that emergency medicine providers also report high professional fulfillment and job satisfaction, suggesting complex relationships between stressors, burnout, and satisfaction in high-acuity environments.^10,12^ While less specialty-specific data exist for urgent care, these clinicians practice in similarly demanding settings characterized by high patient volumes and time pressures.^15^ Taken together, these methodologically distinct but convergent data sources suggest that burnout in emergency and urgent care specialties is increasing rapidly over time and may be underreported, reinforcing the need to examine additional determinants in this high-risk group.

Current burnout prevention strategies typically focus on individual interventions (stress management, mindfulness) or organizational changes (scheduling modifications, wellness programs), yet these demonstrate limited long-term effectiveness.^16–19^

Emerging evidence suggests cognitive mindset may influence how providers experience occupational stressors. Outcome orientation—focusing on desired results rather than problems or blame—has been associated with improved resilience across professional contexts.^20,21^ The Empowerment Dynamic framework proposes that individuals can shift from problem-focused thinking to outcome-oriented patterns emphasizing personal agency.^22^ This aligns with cognitive behavioral theory, which emphasizes cognitive appraisal in determining responses to stressful situations.^23,24^ Preliminary evidence supports this framework: Tetzlaff and colleagues found no difference in oncology PA patient volumes between 2015 and 2019 despite significant burnout increases, suggesting factors beyond workload contribute to distress.^25^

Given these shifts in assessment practices and the evolving understanding of burnout’s multidimensional nature, this study sought to take a comprehensive approach examining relationships between cognitive mindset orientation, burnout, and professional fulfillment among APPs in emergency and urgent care settings. We hypothesized that outcome-oriented patterns would associate with higher fulfillment and lower burnout, while problem-oriented patterns would show opposite associations.

## METHODS

### Study Design and Setting

This cross-sectional study was conducted between July and October 2025 across Advocate Health Organization and Atrium Health Wake Forest Baptist networks. The Advocate Health WFU Health Sciences Institutional Review Board approved this study. A cross-sectional design was selected as appropriate for initial instrument validation and hypothesis-generating research.^26^

### Participants

Eligible participants included licensed APPs (PAs, NPs, Clinical Nurse Specialists) currently practicing in emergency departments or urgent care facilities. Sample size calculations determined 100 participants would provide adequate power (>80%) to detect correlations of r ≥ 0.35.^27,28^

### Recruitment

Participants were identified through administrative databases using census sampling. Survey invitations were distributed via institutional email with up to four reminders.^29^ Surveys were administered through REDCap with quality controls including attention checks and duplicate prevention.^30^

### Measures

The Stanford Professional Fulfillment Index (PFI) is a validated 16-item instrument with subscales for professional fulfillment, work exhaustion, and interpersonal disengagement.^31^ Burnout was defined as average exhaustion and disengagement scores ≥ 3.33; high fulfillment as scores ≥ 7.0.

The Cognitive Mindset and Provider Experience Survey is a newly developed 22-item instrument assessing outcome versus problem orientation based on The Empowerment Dynamic framework.^22^ Instrument development followed established guidelines^32,33^ including literature review, expert panel review by seven multidisciplinary experts, content validity assessment,^34^ cognitive interviews with five practicing APPs for item clarity and comprehension, and pilot testing with 25 providers from non-participating sites to assess response distributions and completion time. Demographics included role, practice setting, experience, employment status, gender, and age.

### Statistical Analysis

Analyses were performed using R version 4.3.2 (α = 0.05). Psychometric validation employed exploratory factor analysis with principal axis factoring and oblique rotation.^35^ Internal consistency was evaluated using Cronbach’s alpha (≥ 0.70 acceptable).^36^ Primary analyses examined Pearson correlations; multivariable linear regression controlled for demographic variables. Missing data were evaluated using Little’s MCAR test.^37^

## RESULTS

### Participants

Ninety-eight APPs completed the survey (19.5% response rate). Respondents included 67 PAs (69.1%) and 30 NPs (30.9%), with majority full-time employment (86.7%) and female gender (82.7%). Practice settings included urgent care (53.6%), emergency departments (29.9%), and both settings (14.4%). Experience ranged from early-career (<3 years, 22.4%) to late-career (>12 years, 35.7%). Variable-level missingness was low (0%–5.1%). Little’s test suggested data were not missing completely at random (P = .04), though the small magnitude indicated no expected bias (Table 1).

**Table 1.** Participant Characteristics (N = 98)

| **Characteristic** | **N (%)** |
| --- | --- |
| **Role** |  |
| Physician Assistant | 67 (69.1) |
| Nurse Practitioner | 30 (30.9) |
| **Practice Setting** |  |
| Emergency Department | 29 (29.9) |
| Urgent Care | 52 (53.6) |
| Both ED/UC | 14 (14.4) |
| Other | 2 (2.1) |
| **Years of Experience** |  |
| <3 years | 22 (22.4) |
| 3–12 years | 41 (41.9) |
| >12 years | 35 (35.7) |
| **Employment Status** |  |
| Full-time | 85 (86.7) |
| Part-time/Per diem | 13 (13.3) |
| **Gender** |  |
| Woman | 81 (82.7) |
| Man | 16 (16.3) |
| Other/Prefer not to say | 1 (1.0) |
| **Age Range** |  |
| 20–39 | 48 (49.0) |
| 40–59 | 47 (48.0) |
| 60+/Prefer not to say | 3 (3.0) |

*Abbreviations: ED, Emergency Department; UC, Urgent Care.*

### Professional Fulfillment and Burnout

Mean professional fulfillment was 5.8 (SD 2.1) on a 0–10 scale, with 38 participants (38.8%) meeting high fulfillment criteria. Mean work exhaustion was 5.2 (SD 2.3), and mean interpersonal disengagement was 3.8 (SD 2.1). Composite burnout was 4.5 (SD 1.9), with 40 participants (40.8%) meeting burnout criteria. Over half (55.1%) did not meet burnout criteria, and 60.2% reported low fulfillment, indicating many APPs experienced neither high fulfillment nor clinical burnout.

### Psychometric Evaluation

Sampling adequacy was strong (KMO = 0.85), and Bartlett’s test confirmed factorability (P < .001). The two-factor solution explained 40% of variance: Factor 1 captured reactive, problem-focused responding; Factor 2 captured self-regulation under pressure. Factors were moderately correlated (r = −0.53). Internal consistency was good (problem orientation α = 0.80; outcome orientation α = 0.89; Supplemental Table).

### Correlational Analyses

Professional fulfillment and burnout were strongly inversely correlated (r = −0.62; 95% CI, −0.73 to −0.48; P < .001). Participants with high fulfillment reported lower burnout (mean 2.9 vs 5.4; mean difference 2.5; 95% CI, 1.9–3.1; P < .001; Cohen d = 1.6). Problem orientation correlated positively with burnout (r = 0.56; P < .001) and negatively with fulfillment (r = −0.36; P < .001). Outcome orientation showed opposite patterns: negative correlation with burnout (r = −0.57; P < .001) and positive with fulfillment (r = 0.44; P < .001). Problem and outcome orientations were inversely correlated (r = −0.63; P < .001) (Table 2, Figure 1). Regression analyses confirmed these associations (Supplemental Figure 1).

**Figure 1.**
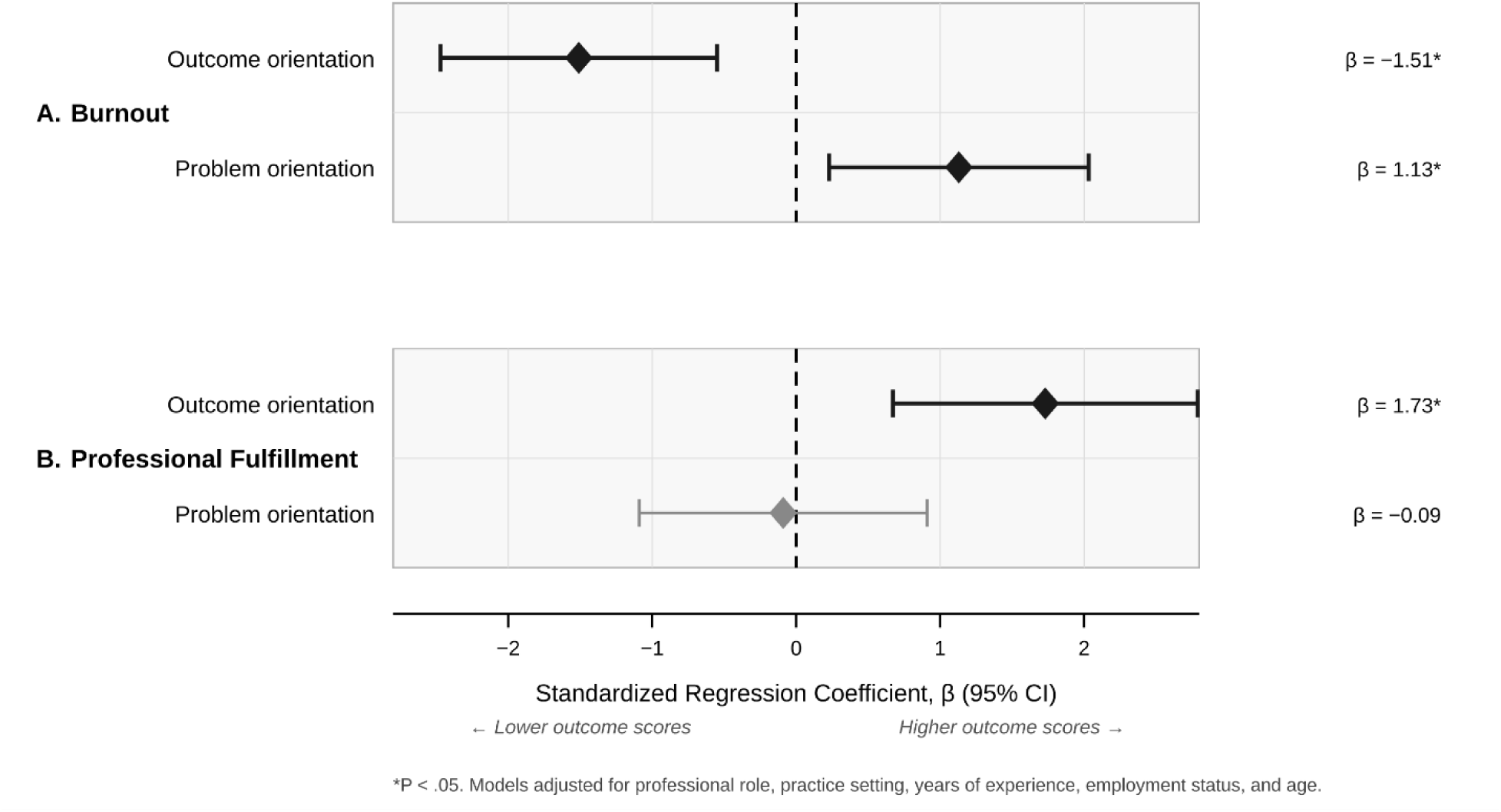
Multivariable Associations Between Cognitive Mindset Orientations and Outcomes *Footnote:* *P < .05. Standardized regression coefficients (β) from multivariable linear models. Models adjusted for professional role, practice setting, years of experience, employment status, and age. Gray diamond indicates nonsignificant association.

**Table 2.** Correlations Between Professional Fulfillment, Burnout, and Cognitive Mindset.

| ***Relationship*** | ***r*** | ***95% CI*** | ***P*** |
| --- | --- | --- | --- |
| Professional fulfillment vs burnout | −0.62 | −0.73 to −0.48 | <.001 |
| Problem orientation vs burnout | 0.56 | 0.40 to 0.69 | <.001 |
| Outcome orientation vs burnout | −0.57 | −0.69 to −0.41 | <.001 |
| Problem orientation vs fulfillment | −0.36 | −0.52 to −0.18 | <.001 |
| Outcome orientation vs fulfillment | 0.44 | 0.27 to 0.59 | <.001 |
| Problem vs outcome orientation | −0.63 | −0.74 to −0.49 | <.001 |

*All correlations are Pearson coefficients (two-tailed tests). All $P < .001$ .*

### Cognitive Mindset Group Comparisons

Participants classified by median splits demonstrated significant burnout differences (F = 11.8; P < .001). Outcome-oriented participants had lowest burnout (mean 3.2; SD 1.4), while problem-oriented participants had highest (mean 5.8; SD 1.7; mean difference 2.6; 95% CI, 1.5–3.7; P < .001). Fulfillment also differed significantly (F = 2.7; P = .048), with outcome-oriented participants reporting highest fulfillment (mean 6.8; SD 1.9) and problem-oriented lowest (mean 4.7; SD 2.0) (Figure 2).

**Figure 2.**
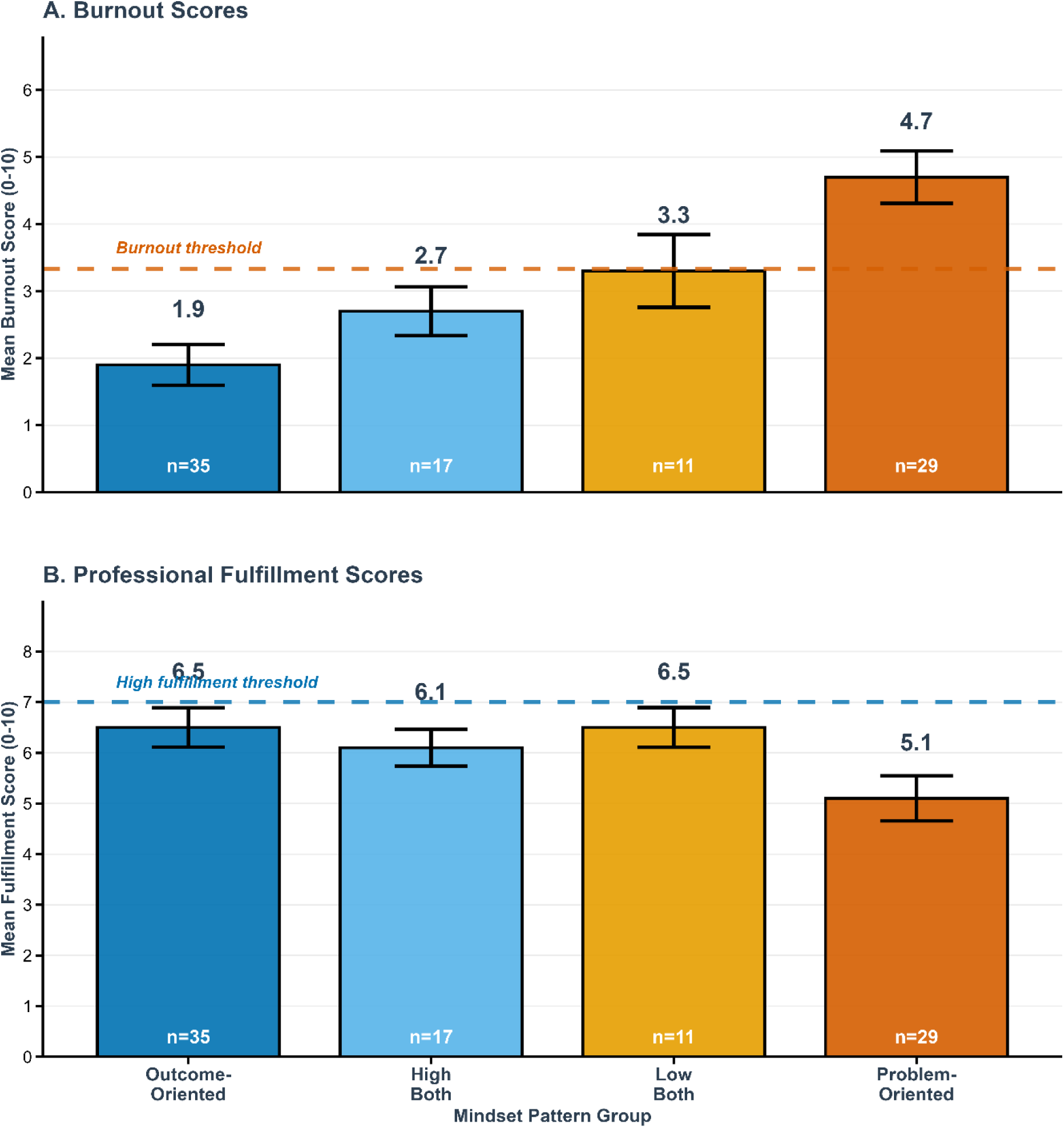
Burnout and Professional Fulfillment by Cognitive Mindset Pattern *Footnote:* Burnout and professional fulfillment scores by cognitive mindset pattern. Participants classified into four groups by median splits: Outcome-Oriented (low problem/high outcome), Problem-Oriented (high problem/low outcome), High-Both, and Low-Both. Error bars represent ±1 standard error. Burnout threshold = 3.3; high fulfillment threshold = 7.0. Between-group differences significant for burnout (F = 11.78, P < .001) and fulfillment (F = 2.74, P = .048). Pairwise differences tested using Tukey HSD.

### Multivariable Analyses

After adjusting for role, practice setting, experience, employment status, and age, both orientations independently predicted burnout: problem orientation positively (β = 1.13; P = .015) and outcome orientation negatively (β = −1.51; P = .003), explaining 35.6% of variance. For fulfillment, outcome orientation remained significant (β = 1.73; P = .002), while problem orientation was not (β = −0.09; P = .86). No significant differences emerged by professional role or practice setting.

## DISCUSSION

This study examined relationships between cognitive mindset orientation, professional fulfillment, and burnout among APPs while providing preliminary validation of a novel assessment instrument. Outcome-oriented cognitive patterns were associated with significantly lower burnout and higher professional fulfillment, independent of demographic and practice characteristics.

The 40.8% burnout prevalence aligns with recent national estimates for emergency medicine and urgent care PAs.^12^ However, 60.2% of participants reported low fulfillment without meeting burnout criteria, highlighting that absence of burnout differs from presence of professional satisfaction—a distinction often overlooked in workforce research.^38,39^ This suggests interventions must both reduce negative experiences and cultivate positive ones.

Problem and outcome orientations were strongly inversely correlated (r = −0.63), supporting their conceptualization as distinct cognitive patterns.^22^ Critically, multivariable regression revealed both independently predicted burnout, with outcome orientation uniquely predicting fulfillment. This suggests promoting outcome-oriented thinking may be more effective than simply reducing problem-focused cognition.

Our findings extend research showing workload inadequately explains burnout variability.^25,40^ The correlations between cognitive orientation and well-being (r = 0.36–0.57) are comparable to effect sizes for emotional intelligence, self-compassion, and psychological capital in healthcare workers.^41–43^ However, cognitive orientation may be more modifiable than temperamental factors, making it a promising intervention target.^44^

### Clinical and Research Implications

If validated prospectively, cognitive mindset assessment could identify at-risk APPs before symptoms develop. Intervention development could incorporate cognitive behavioral therapy techniques, which have reduced physician burnout in randomized trials.^45,46^ However, individual-level screening must be embedded within larger organizational efforts. Healthcare systems and enterprises bear responsibility for proactive assessment, recognition, and support of their workforce.^1,47^ Understanding the relationship between burnout, fulfillment, and cognitive patterns can inform system-level responses required to maintain a healthy, high-functioning healthcare workforce. Screening approaches must avoid stigmatizing individuals or shifting responsibility for systemic problems onto providers.^48^

### Limitations

The cross-sectional design precludes causal inference; longitudinal studies are needed to clarify temporal relationships.^49^ Our sample of 98 participants from two health systems limits generalizability. The predominantly female sample (82.7%), while reflecting APP workforce demographics,^50^ raises considerations regarding gender differences in self-reporting practices, as research suggests potential variations in willingness to disclose emotional experiences.^51^ The 19.5% response rate raises selection bias concerns, as clinicians most affected by burnout may have been least likely to participate.^52^

The cognitive instrument requires comprehensive validation with larger samples and confirmatory factor analysis.^53^ Self-report measures introduce common method variance and potential under- or over-reporting of emotional states.^54^ We did not directly assess organizational variables known to influence burnout, preventing examination of interactions between cognitive orientation and environmental stressors.^55,56^

Critically, individual cognitive factors must be understood within broader systemic determinants. The National Academy of Medicine emphasizes individual interventions alone are insufficient—meaningful burnout reduction requires organizational changes addressing workload, autonomy, and work-life integration.^1^ Cognitive orientation should not be viewed as causing burnout or as the sole solution, but as one potentially modifiable factor within a complex system.

### Future Directions

Longitudinal cohort studies should examine whether baseline cognitive orientation predicts burnout incidence and recovery. Randomized controlled trials testing cognitive interventions versus active controls would establish efficacy.^45^ Given the variability in how national bodies assess and report burnout, future research would benefit from triangulated approaches using multiple validated measures to enhance comparability across studies.^11,12^ Furthermore, healthcare systems should consider taking ownership of regular assessment and recognition of provider well-being, moving beyond reliance on external surveys toward integrated organizational monitoring and support structures.^47,57^

## CONCLUSIONS

This study provides preliminary evidence that cognitive mindset orientation is associated with professional fulfillment and burnout among APPs in emergency and urgent care settings. The novel assessment instrument demonstrated acceptable psychometric properties warranting further validation. Cognitive factors represent potentially modifiable elements influencing provider well-being, though should complement—not replace—organizational interventions. Future longitudinal studies and intervention trials are needed to establish causality and clinical utility.

## Ethics Statement

This study was approved by the Advocate Health WFU Health Sciences Institutional Review Board. Informed consent was obtained from all participants prior to survey completion.

## Data Availability

All data produced in the present study are available upon reasonable request to the authors

## Acknowledgments

The authors thank the Advanced Practice Providers who participated in this study. This manuscript was created as part of graduation requirements for the DMSc program of Wake Forest University School of Medicine Physician Assistant Studies. A pre-press copy of this manuscript has been posted to https://www.medrxiv.org

